# Understanding soaring coronavirus cases and the effect of contagion policies in the UK

**DOI:** 10.1101/2021.01.30.21250822

**Authors:** Miguel A. Durán-Olivencia, Serafim Kalliadasis

## Abstract

The number of new daily SARS-CoV-2 infections is frantically rising in almost every country of the EU. The phenomenological explanation offered is a new mutation of the virus, first identified in the UK. We use publicly available data in combination with a controlled SIR model, which captures the effects of preventive measures on the active cases, to show that the current wave of infections is consistent with a single transmission rate. This suggests that the new SARS-CoV-2 variant is as transmissible as previous strains. Our findings indicate that the relaxation of preventive measures is closely related with the ongoing surge in cases. We simulate the effects of new restrictions and vaccination campaigns in 2021, demonstrating that lockdown policies are not fully effective to flatten the curve. For effective mitigation, it is critical that the public keeps on high alert until vaccination reaches a critical threshold.

---

The susceptible-infectious-recovered (SIR) model (1) (Fig. 1.a), is a popular vanilla model for computational scrutiny used in numerous studies, often to estimate the characteristic transmission rate of SARS-CoV-2. However, despite its flexibility and mathematical elegance, the model introduces some important limitations. To determine whether new variants of the SARS-CoV-2 are more transmissible than their predecessors, data analysis must cover the entire pandemic outbreak and include the effects of preventive measures and contagion policies taken by populations and governments, respectively. The model also does not account for the social preventive response which characterises the *new normal*, e.g. social distancing, mask wearing, limited commuting, remote working, or local curfews and lockdowns, to name but a few examples. Fitting data to a model which does not capture how these important social changes affect the spreading of the virus is not a reliable method to discern the transmission (*β*), recovery (*α*) and basic reproductive 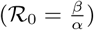 rates. For instance, different values of ℛ_0_ for the same virus with the same inherent properties under different social contexts, e.g. partial and full lockdown, are obtained. Data for the United Kingdom suggest much higher values for *β* and ℛ_0_ during September-December 2020 if analysed under the SIR premises (2). Despite the good fitting of data achieved over limited temporal windows (2–5), there are two important limitations compromising the accuracy of the predictions: a) one cannot fit the whole temporal series, characterised by multiple infection waves, and indeed the fit would eventually diverge; and b) the SIR model, or the equivalent logistic growth model (2), would never forecast a second or further upsurge in cases. Refined SIR models to include additional factors, such as “shield immunity” (6), do not come to our rescue. Indeed, despite their increased flexibility they are still only capable of showing a single wave. Additionally, many studies fitted SIR-like models to data from the last stages of the first wave – even though the models suffer from the inherent limitation of a single-wave prediction – thus effectively assuming that the epidemic was coming to an end. Yet it was already known at the time that the number of cases was decreasing because of the preventive measures which in turn should have been sufficient to abandon the corresponding models. Thus, current studies are incomplete and an alternative approach is necessary.

**Fig 1.**
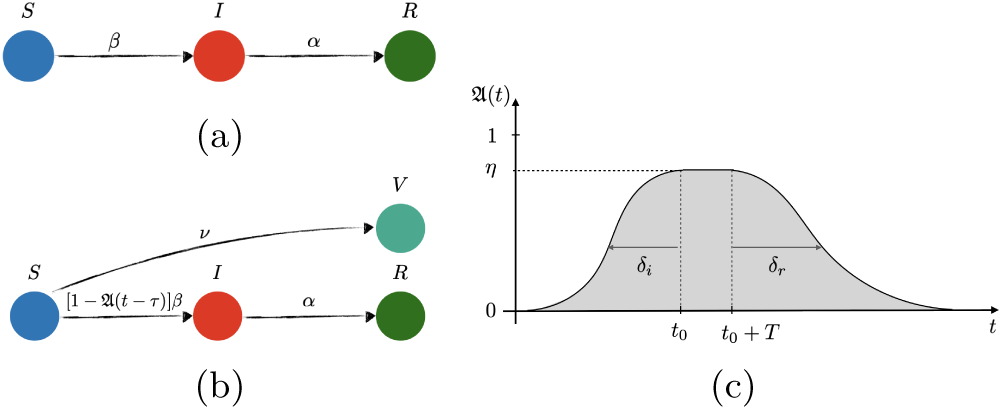
Sketch of transitions in (a) free and (b) controlled SIR network model of disease transmission, and (c) preventive social response, 𝔄(*t*).

The main results of the present work are as follows. Consideration of the full-history of the data with a *controlled* SIR model (Fig. 1.b) avoids the drawbacks of previous models, by capturing the essence of how the new normal affects the number of infected people. This unveils unique and constant *β* and ℛ_0_ for the entire pandemic. Thousands of mutations have emerged in the SARS-CoV-2 genome since the first outbreak in 2019, and only the UK strain, known as B.1.1.7, is being reported as a more “aggressive” form of the virus, because of an alarming surge in new cases thought to be correlated with the new UK variant, and was one the reasons for the lockdown imposed in the UK at the beginning of 2021, e.g. Ref. (7). According to the law of parsimony: chose the simplest explanation from those that fit. Indeed, our results show that the fierce increase in cases is captured without the need of a more transmissible variant, suggesting that genomic data during the pandemic might have been overinterpreted. Our approach includes characteristic parameters which could be pivotal in the decision-making process in the coming months. For instance, there seems to be an *inertia of society* which plays a crucial role on the flattening of the curve. For preventive measures to be effective, these should be encouraged quite early in the surge of cases, taking into consideration the inherent social inertia, which typically leads up to three-weak delay up until society gets to its maximum level of alert. We also include the effect of vaccination, and show that *social relaxation* as of March 2021 without fulfilling a sufficient vaccination rate(determined below) will lead to a new wave of infections over May-June 2021, independently of the more strict lockdown currently imposed since January 2021.

Figure 2 reports curve fits and predictions of the free and controlled SIR models. The free version (Fig. 1.a) fits well the data of the first wave of infections from March to June 2020, but completely fails to predict any second or further wave. This is because in a free SIR model the decay of the infected cases is only possible when the pandemic is already in recession. [As we know now and back in June 2020, this was not the reason for the decrease in cases back then; rather the reduction of susceptible people is due to preventive measures.] Yet has been a standard way of extracting estimates for *β, α* and ℛ_0_. Some works even published estimates for these quantities via manual fitting to data (5), effectively a trial-and-error approach. The authors justified this by asserting that a rigorous non-lineal fitting did not follow the data as close as their approach.

**Fig 2.**
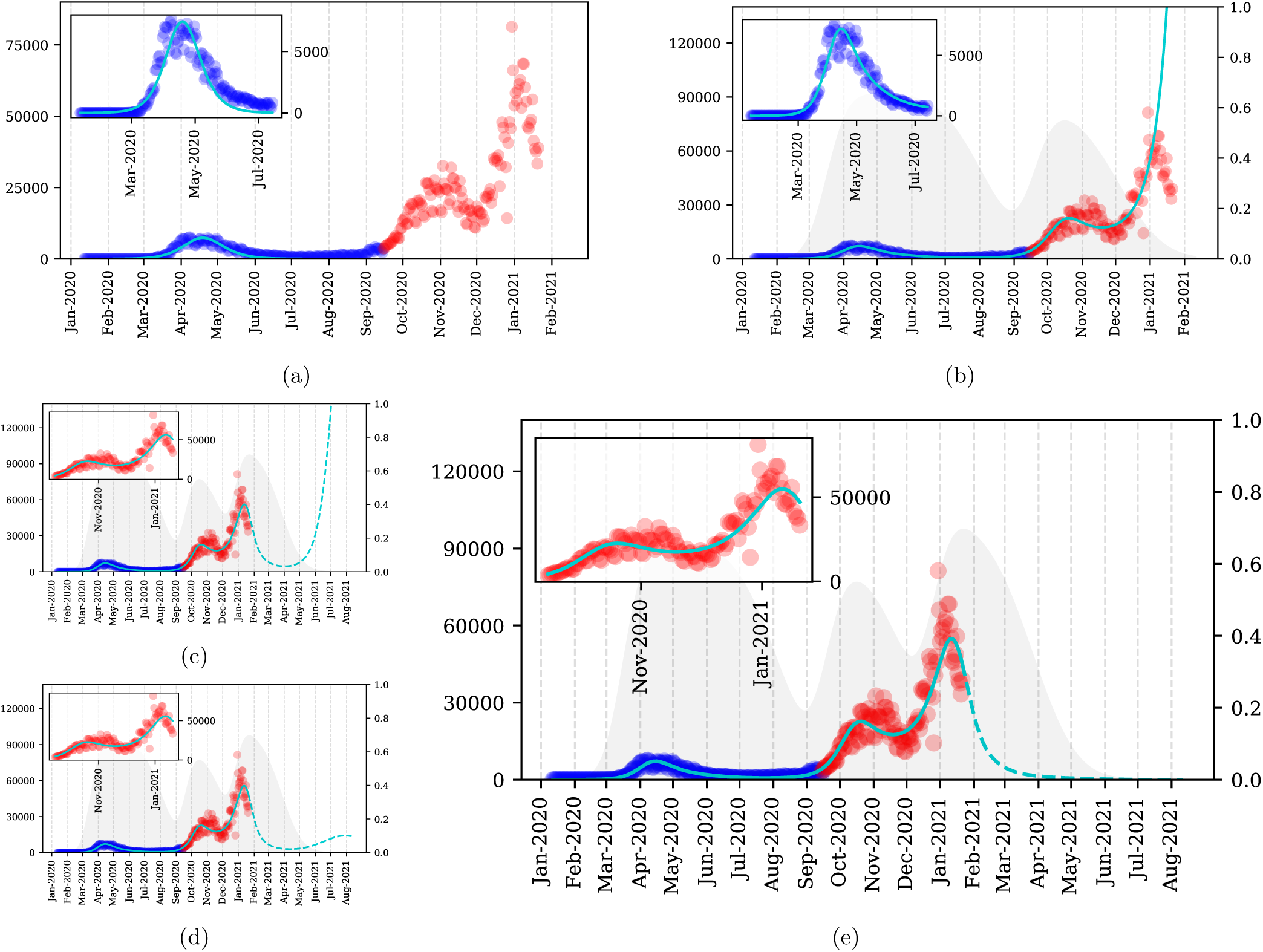
SARS-CoV-2 new daily cases (left axis): blue circles for first-wave data used to fit free and controlled SIR models (light-blue lines), red circles for second- and third-wave data used to test the models and their predictions (light-blue dashed lines). (a) Free SIR model captures the essence of the time evolution of new CoVid-19 cases over March-July 2020 (inset plot), but totally fails to predict the second and third waves. (b) Controlled SIR model without vaccination fits better to first-wave data (inset plot) than the free version. The gray area represents the effectiveness of preventive measures, 𝔄 (*t*) (right axis). The first wave of social awareness is fit together with *β* and *α*, showing a maximum of effectiveness *η*_1_ ≃65%, social inertia *δ*_*i*_ ≃21 d, and social relaxation starting at mid June 2020, with prediction of no measures in *δ*_*r*_ ≃45 d after relaxation begins. The second wave of social awareness begins in September (confirmed by Prime Minister (9)), reaching *η*_2_ ≃60% by mid October 2020 (three-tier system was introduced (10)). The upsurge of CoVid-19 cases in December 2020 is again a consequence of social relaxation. (c)-(e) Controlled SIR model with vaccination rates: 0.1%d^−1^, 0.2%d^−1^, and 0.4%d^−1^, respectively, along with a third-wave of preventive measures (expected to reach maximum effectiveness, *η*_3_ = 70%, by the mid January 2021). To avoid a fourth wave, the vaccination campaign would need to deliver ∼ 200 × 10^3^ vaccines per day (∼ 0.4%*N/*d) as of the first week of January 2021.

**Fig 3.**
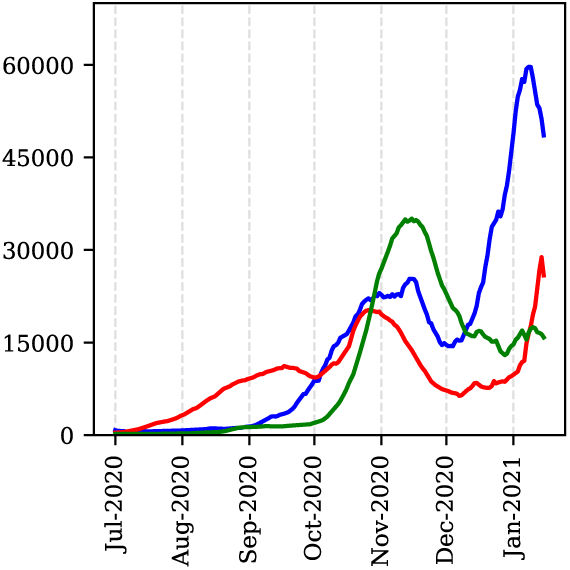
Daily new cases (7-day moving average): UK (blue), Spain (red) and Italy (green).

However, instead of imposing the SIR model and changing the fitting method to achieve agreement with the model, the disagreement with a nonlinear fit means that they should have abandoned the model.

Not only does the controlled SIR model fits better the first-wave data (inset plot, Fig. 2.b), but also captures the reason for the decrease in cases from mid April to August 2020, namely a wave of *social awareness* (A) which effectively reduced the number of susceptible people. 𝔄 embodies both contagion policies and the citizens efforts made to “flatten the curve”, e.g. wearing masks, reducing travelling or self-isolating. Moreover, the model predicts a sudden rise in cases when society relaxes, because the downtrend in new infections is not related with the end of the pandemic but with a temporary removal of susceptible candidates from the system. This is precisely what happened from July to September 2020, and what eventually led to the surge in cases in early September 2020. This sharp increase immediately raised the alarm (8, 9), and 𝔄 started growing again, reaching a maximum effectiveness when the three-tier restrictions system was imposed (10). However, these measures were not enough to flatten the curve and a new increase appeared in December 2020 because of a gradual relaxation over the month of November. By incorporating new waves of preventive measures in 𝔄 the model is able to reproduce the above observations, as illustrated in Figs 2.b-e. This provides evidence of the predictive capabilities of the model.

Figures 2.c-e reveal the effects of the third lockdown imposed on January 4, 2021 and of the vaccination campaign with rates 0.1%d^−1^, 0.2%d^−1^, and 0.4%d^−1^, respectively. The trends illustrate that unless the campaign delivers 200 10^3^ vaccines per day, a fourth wave is unavoidable. At the same time, if the vaccination rate is less than 100 × 10^3^ vaccines per day, the fourth wave will be as severe as previous ones.

Analysis of European countries’ data depicted in Fig.3, including countries severely hit by the pandemic, like Spain and Italy, reveals that virtually in all cases, the UK pattern persists and similar conclusions can be drawn. This means, social relaxation – typically two-to-three months – driving an abrupt increase of cases, followed by increased awareness and preventive measures leading to temporal stagnation, which is then followed by further growth, i.e. a third, more powerful, wave.

## Methods

### Population dynamics

The population is split into four groups: susceptible (*S*), infected (*I*), recovered (*R*) and vaccinated (*V*), as illustrated in Fig.1. The groups follow the delayed dynamical system:

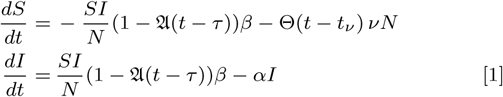

where 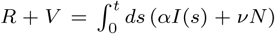, *N* = *S* + *I* + *R* + *V* is the total population, assumed 60 million. The parameters *β, α, ν* are the transmission, recovery, and vaccination rates, respectively, and 𝔄 (*t* − *τ*) is the percentage of susceptible people using effective preventive measures at time *t* − *τ*, (*τ* being a characteristic time for such preventive measures to become apparent, assumed 14 days, the incubation period,(14)) with general functional form: 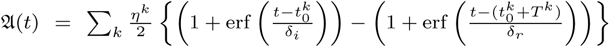 where *η*^*k*^ ∈ [0, 1] is the effectiveness of the preventive measures taken in the *k* th wave, *T* ^*k*^ is related to the time extension of these measures, and *δ*_*i,r*_ are the social inertia (*i*) and relaxation (*r*) time scales, respectively. With 𝔄 = 0 and *ν* = 0, Eq. (1) becomes the free SIR model. For 𝔄 ≠ 0 we get a controlled SIR model. The initial condition used: *I*_0_ = 1 (number of infective cases reported on January 11, 2020), *S*_0_ = *N* − *I*_0_ and *R*_0_ = *V*_0_ = 0. Thus fitting of five parameters is needed, and this is done at the first wave.

### Training and testing the model

The full dataset, *Y*, was renormalized 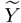, by using a linear fit, *z* = *b* + *mx*, to the number of daily CoVid-19 tests per thousand people given in Ref. (11), with *b* = 0 and *m* = 0.0191 test*/*103people*/*d (from 0 to 7 test*/*103people in 366 days), so that 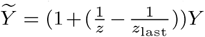. We then use non-linear least squares to fit the daily new cases 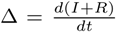 to the first-wave data (training dataset). This fitting yields for the free model: *β* = 0.515d−1 and *α* = 0.420d−1 so that 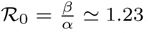. For the controlled model we get: *β* = 0.211 d−1, *α* = 0.102 d−1, *η* = 0.65, 140 d, *δ*_*a*_ = 21 d and *δ*_*r*_ = 45 d, which yields ℛ_0_ ≃2.068. To test the models we numerically integrate Eq. (1) for both cases, i.e., 𝔄 (*t*) = 0 and 𝔄 (*t*) 0. The controlled-model prediction for new infections grows exponentially as of September 2020 (testing dataset) when the first wave of preventive measures would vanish according to the summer trend. With *δ*_*i,r*_ fixed from the first wave, we fit the parameters *η* and *T* of a second social response to unveil the behavioural changes adopted against the apparent second wave of cases, obtaining a maximum of social response by mid-end October 2020. This is in perfect agreement with the declaration of the UK Prime Minister “seeing a second wave” on September 18, 2020 (9), and his statement on coronavirus where the three-tier restrictions system was imposed on October 12, 2020 (10). For predictions as of January 2021, we introduce a third wave of measures aiming at an 70% of effectiveness, i.e. *η* = 0.70, starting in January and ending in April 2021, *T* = 90 d, which represents the current contagion policies being taken by the UK government. Finally, we adopt three potential values for 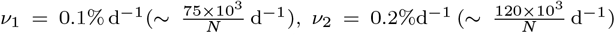 and 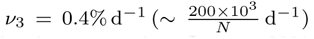, with *ν*_3_ being the current vaccination target since January 2021 (13).

## Data Availability

All data for the analysis was collected from https://www.worldometers.info/coronavirus/country/uk/

## Data Availability

All data for the analysis was collected from https://www.worldometers.info

## ACKNOWLEDGMENTS

We acknowledge financial support from the Engineering and Physical Sciences Research Council of the UK via grant No. EP/L020564/1.

